# Effects of multimodal and unimodal physical training interventions on visual function in glaucoma and elderly controls – a pilot study

**DOI:** 10.1101/2025.01.30.25321400

**Authors:** DC Moffack, KO Al-Nosairy, R Beyer, C Freitag, FH Stolle, M Behrens, T Behrendt, GT Prabhakaran, H Thieme, L Schega, MB Hoffmann

## Abstract

**Objective:** To investigate the effect of a 12-week multimodal motor-cognitive and resistance training (MMI) compared to a unimodal resistance training intervention (UMI) on static and dynamic visual function in glaucoma patients (GLA) and healthy controls (HC).

**Methods:** Fifteen GLA and 24 age-matched HC participated in this randomized, controlled longitudinal pilot study. Visual function was assessed using clinical parameters and, on a treadmill, (TM) while standing (static, S_0_) as well as during walking at 3.5 km/h (S_3.5_) and a self-preferred speed (S_Self_). The following outcomes were measured pre and post 12-weeks of intervention (MMI or UMI): (a) standard clinical measures and (b) TM-related measures, i.e. (i) best-corrected visual acuity without (VA_S_) and with crowding (VA_C_), (ii) visual field sensitivity (VF), and (iii) contrast sensitivity (CS). A 4-factorial repeated-measures ANOVA (SPEED [S_0_; S_3.5_; S_Self_] x TIME [pre; post] x INTERVENTION [UMI; MMI] x GROUP [GLA; HC]) was applied to determine significant interaction effects (p ≤ 0.05) and the effect size partial-eta-squared was calculated.

**Results:** Post-interventional improvement of visual function was absent or minor. Only for standard clinical measures a main effect of TIME (visual acuity; p=0.024) and a TIME x INTERVENTION interaction (foveal sensitivity; p=0.039) were found. For S_0_ vs S_3.5_ small effects appeared in post-hoc comparisons, but TIME and TIME x SPEED just failed to reach significance for CS (p=0.059) and VA_s_ (p=0.052), respectively.

**Conclusion:** While trends were evident, the effect of the 12-week interventions on visual function were small and, especially for TM-walking, largely independent of group and intervention type. In future studies a greater sample of more advanced glaucoma cases should be included to probe for significant visual function differences between groups and intervention types.

## Introduction

Glaucoma, a progressive optic neuropathy, is a leading global cause of irreversible blindness. Current estimates expect an increase of affected individuals from 76 million to 112 million by 2040 (Tham et al., 2014). Glaucoma damage impedes activities that are of importance for daily living not exclusively in the visual domain, but also with respect to mobility, e.g. via an increased incidence of falling during walking (Freitag et al., 2023). As a consequence, glaucoma has an impact at the individual level due to quality of life reduction (Fea et al., 2017; Moreno-Montañés et al., 2018) and at the societal level due to health-care cost increase (Allison et al., n.d.; Feldman et al., 2020). However, these conventional measures often overlook a broader aspect of functional impairment in glaucoma patients, particularly those related to motor and cognitive abilities (Tanabe et al., 2012). It has been shown that resistance training mitigated the risk of falls in elderly (Fragala et al., 2019; Persch et al., 2009) and might also increase cognitive performance (Fiatarone Singh et al., 2014). Recent reports (Herold et al., 2018) also indicated that the combination of motor and cognitive exercise has beneficial effects on motor and/or cognitive function in patients with neurodegenerative diseases, e.g., Alzheimer’s disease ( Coelho et al., 2013; Puente-González et al., 2021) and in healthy participants with cognitive decline (de Oliveira Silva et al., 2019). Moreover, this interventional approach might reduce the risk of falls (Fragala et al., 2019; Persch et al., 2009).

Due to the fact that glaucoma might be associated with cognitive dysfunction (Arrigo et al., 2021), combining motor-cognitive exercises (Herold et al., 2018) with resistance training in glaucoma management might be of promise to improve mobility and cognitive function eventually translating in an increased quality of life. This prompts the question of whether a combination of motor-cognitive and resistance training (i.e., multimodal intervention [MMI]) is superior in improving motor-cognitive function compared to resistance training alone (i.e., unimodal intervention [UMI]) in glaucoma patients, which has, to our knowledge, not been addressed so far.

The present study reflects on a subset of data from a larger randomized controlled trial (German clinical trials register: DRKS00022519) examining the effects of two physical training interventions (MMI and UMI) on several primary and secondary outcomes. The primary endpoints include gait kinematics and functional brain connectivity.

In the present pilot study, visual function, as a secondary outcome, is addressed. Specifically, we investigated the influence of a 12-week MMI and a UMI on visual function during treadmill (TM) walking in glaucoma patients and healthy elderly controls. The baseline data, i.e. pre-intervention findings, of this study were previously published (Beyer et al., 2024) and indicated that TM walking, compared to standing, reduced visual function similarly in both glaucoma patients and elderly controls (i.e. a visual acuity loss by more than 0.02 logMAR and visual field sensitivity by 1.0 dB visual field mean deviation). Consequently, it is of interest whether MMI or UMI can improve visual function and thus ultimately, reduce the risk of falls. We hypothesized that visual function will be improved after the interventions in glaucoma, especially following MMI, as this intervention specifically targets visuo-cognition compared to UMI.

## Methods

### Study design

To investigate the effect of MMI and UMI, a two-arm randomized controlled prospective longitudinal study was conducted from August 2020 to December 2022. Patients with glaucoma (GLA) and age-matched healthy control subjects (HC) were recruited and randomly assigned to either MMI or UMI using counterbalanced randomization and allocation ration of 1:1 (Figure 1).

**Figure 1.**
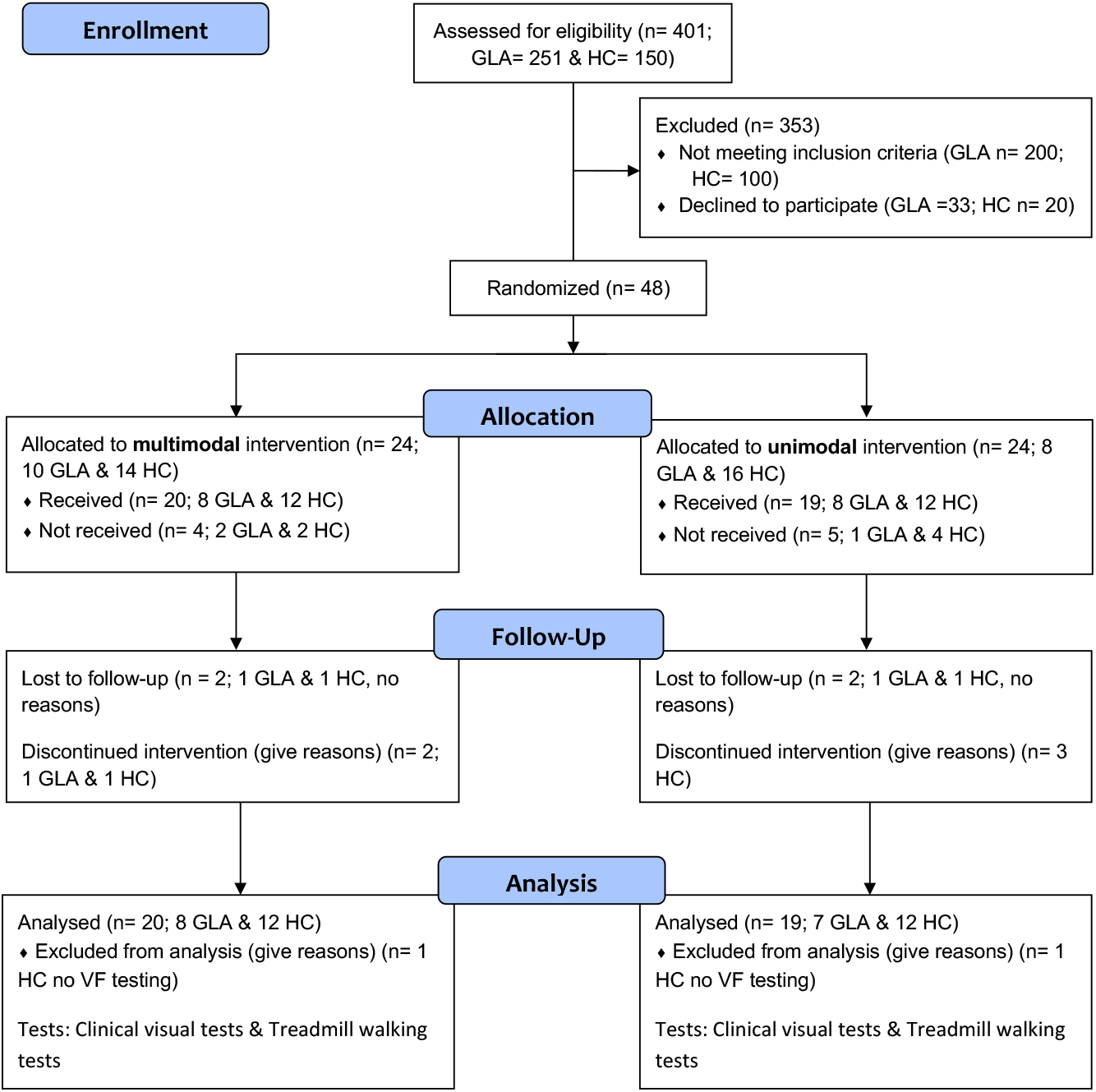
CONSORT flow chart for the recruitment process showing the number of participants and dropouts through the study phases. GLA = glaucoma participants; HC = healthy participants

The experimental protocol was approved by the Ethics Committee of the Otto-von-Guericke University of Magdeburg in Germany (registration number: 32/18) and all procedures were in line with the Declaration of Helsinki on experiments on human beings. All visual-function related measurements were taken in the Department of Ophthalmology and at the Otto-von-Guericke University Magdeburg, Germany. The interventions were performed in the Department of Sport Science at the Otto-von-Guericke University Magdeburg, Germany. Reporting was performed in accordance with the Consolidated Standards of Reporting Trials Statement (Consort) for randomized pilot trials (Eldridge et al., 2016). All participants gave written informed consent. The trial was registered at the German clinical trials register (DRKS00022519).

### Participants

A total of 39 participants, including 15 GLA and 24 HC, were recruited in this study. For GLA, only two of the participants had an advanced stage, two had a mild stage and the rest had a preperimetric stage.

Participants underwent complete ophthalmological examination, which is described in detail by (Freitag et al., 2024). Briefly, we assessed best corrected visual acuity, refractive correction for far (at 5 m) with early treatment diabetic retinopathy study charts (ETDRS), slit-lamp exam, fundus exam, and standard tests for visual field (VF) and retinal structure. The following inclusion criteria were applied: (i) ≥ 60 years, (ii) diagnosis of open-angle glaucoma (i.e., cup disc ratio of ≥ 0.7, disc notching, and/or nerve fiber defect), (iii) patients are under intraocular pressure medications, (iv) best corrected visual acuity (BCVA) ≥ 0.8 decimal unless glaucoma-related in the GLA group, (v) normal visual function parameters unless glaucoma related, (vi) no other conditions affecting visual function such as age-related macular degeneration or retinal detachment, (vii) ability to walk at least 6 min without walking support, and (viii) visual field defects classified according to Hodapp-parish Anderson criteria (Hodapp et al., 1993) in GLA with defects. Exclusion criteria were as follows: i) neurological disorders, (ii) rheumatism, (iii) cardiovascular disorders, (iv) stroke, (v) orthopedic diseases including arthrosis (grade II or higher), musculoskeletal impairment, tendinitis, tenosynovitis, myositis, prosthesis in the lower extremities, and joint replacements.

### Interventions

The training (i.e., MMI and UMI) was conducted twice a week on non-consecutive days for 12 weeks resulting in a total of 24 sessions. Each session lasted 60 min and was supervised by experienced instructors.

The MMI was split into i) motor-cognitive training, ii) resistance training, and iii) cool-down (i.e., static stretching). Over the course of the intervention, the proportion of time spent on motor-cognitive and resistance training changed (see Table 1). The motor-cognitive exercises consisted of simultaneously performed motor, cognitive, and visual tasks based on the LifeKinetik program (Lutz, 2021). The exercises were designed in such a way that it is almost impossible to perform them without making mistakes. If the exercises were performed correctly in 6 out of 10 attempts, the instructor continued with a more difficult exercise. As examples, two exercises are briefly explained below: (i) balls of different colors (e.g. yellow, green, red) are thrown in a circle, whereby a corresponding name must be said for each color (e.g, yellow = persons own name, green = name of the person to whom the ball is to be thrown. (ii) participants line up next to each other and after an announcement (e.g., left, right, front, back) they walked in the corresponding direction (line of vision remained the same, i.e. no returns), whereby a corresponding name for each direction were varied (e.g. right = “1”, left = “2”, front = “3”, back = “4”).

**Table 1.** Multimodal training schedule.

| Training components | Course of Intervention |  |  |
| --- | --- | --- | --- |
|  | Week 1-4 | Week 5-8 | Week 9-12 |
| Motor-Cognitive Training | 10 min | 20 min | 30 min |
| Resistance Training | 40 min | 30 min | 20 min |
| Cool Down | 10 min | 10 min | 10 min |

The UMI consisted of i) 10-min standardized warm-up (i.e., 5 min fast walking and dynamic stretching), ii) 40-min resistance exercises, and iii) 10-min cool-down (i.e., static stretching). The resistance training included multiple-set (2 x 7 repetitions, 30 s rest between sets, time under tension: 2 s concentric and eccentric) circuit training using free weights and weight machines (e.g., dumbbell seated front raise and seated leg press) (Table 2). External training load was adjusted by increasing the weight and controlled via individuals’ rating of perceived exertion (RPE) using the Borg CR 10 Scale (i.e., RPE = 3-4 (moderate to somewhat severe)).

**Table 2.** Resistance exercises.

| Week | UMI | MMI |
| --- | --- | --- |
| **Weeks 1 – 4** | 1. Seated leg press<br>2. Lever pulldown<br>3. Lever seated twist<br>4. Lever chest press<br>5. Cable curl<br>6. Lever back extension<br>7. Cable pushdown<br>8. Cable upright row<br>9. Seated leg curl | 1. Seated leg press<br>2. Lever pulldown<br>3. Lever seated twist<br>4. Lever chest press<br>5. Cable curl<br>6. Lever back extension<br>7. Cable pushdown<br>8. Cable upright row<br>9. Seated leg curl |
| **Weeks 5 – 8** | 1. Seated leg press<br>2. Cable straight back seated row<br>3. Lever seated twist<br>4. Lever chest press<br>5. Cable curl<br>6. Lever back extension<br>7. Cable pushdown<br>8. Cable upright row<br>9. Lever leg extension | 1. Seated leg press<br>2. Cable straight back seated row<br>3. Lever seated twist<br>4. Lever pec deck fly<br>5. Cable upright row<br>6. Cable curl |
| **Weeks 9 – 12** | 1. Seated leg press<br>2. Cable straight back seated row<br>3. Lever chest press<br>4. Lever seated twist<br>5. Reverse cable curl<br>6. Dumbbell seated front raise<br>7. Seated leg curl<br>8. Lever back extension<br>9. Cable pushdown | 1. Seated leg press<br>2. Dumbbell seated front raise<br>3. Reverse cable curl<br>4. Lever chest press<br>5. Lever seated twist |

Resistance training was carried out in the same way as the MMI, although the number of exercises and its duration varied (see Table 1 and Table 2). All exercises were performed while sitting because exercising in a supine or other positions affects the intraocular pressure (Al-Nosairy et al., 2020; Lara et al., 2023). Both interventions ended with a 10-min cool-down consisting of static stretching for the major muscle groups (e.g., standing side stretch, standing forward fold, overhead triceps stretch).

### Outcome parameters

At the start of this study, participants’ demographic and anthropometric data as well as their physical activity level, and visual function were recorded. Standard ophthalmology tests included: i) standard automated perimetry (SAP) for visual field estimation with 24-2 SITA-Fast test (Humphrey Field Analyzer, Carl Zeiss Meditec AG, Jena, Germany) and ii) optical coherence tomography (OCT) of macula and disc using a spectral-domain OCT device with Glaucoma Module Premium Edition (Heidelberg Spectralis®, Heidelberg Engineering, Heidelberg, Germany). Of note, single- and dual-task gait performance (i.e. stride length, gait velocity) as well as gaze behavior (i.e. saccade duration, fixation duration) were also assessed at the beginning of the study (Freitag et al., 2024, 2023). Effects of the interventions were scrutinized for (A) the standard clinical tests including BCVA, VF parameters, e.g., foveal sensitivity and VF MD and for (B) TM walking testing. For TM walking testing, four visual function tests were performed binocularly [BCVA without crowding (VA_s_), BCVA with crowding (VA_c_), contrast sensitivity (CS) and binocular visual field (Bin-VF)] as detailed below.

### Visual function during TM walking

Visual function was tested at 5 m and to account for variable viewing distance during TM walking as a confound factor, visual functions were corrected using a distance sensor (Vivior® sensor) as detailed in Beyer et al. 2024.

### TM walking testing procedure

Visual functions were tested for 3 speed conditions in a fixed order: i) static (S_0_); ii) TM speed of 3.5 km/h (S_3.5_) and iii) a self-preferred TM speed (S_Self_).

### Visual function testing procedure

For each speed condition, three binocular visual tests were performed respectively using Freiburg Vision Test (FrACT®) (Bach, 1996) in a dimly-lit room at 5-m distance and repeated twice in the following order: i) VA_S_ ii) VA_c_, and iii) CS. This was followed by one Bin-VF test using Ocusweep (Ocusweep^®^, Ocuspecto Ltd, Turku, Finland) in an ambient room light. Total experiments duration was about 75 min.

1. VA_s_: An 8-alternative-forced choice (AFC) single Landolt ring I was presented to estimate VA and reported as a logarithmized minimal angle of resolution (logMAR).
2. VA_c_: The same applies to VA_C_ but with using a single optotype surrounded by a circle (‘©’).
3. CS: An 8-alternative-forced choice (AFC) optotype with varying contrasts was used to determine the weber contrast (logCS).
4. Bin-VF: A square stimulus comprised of 9 LEDs (5.2 mm x 5.2 mm) was presented at near distance (∼40 cm) without proximity sensor to estimate the mean deviation of VF sensitivity (MD). Ocusweep^®^ allowed for VF testing while TM walking since it requires no fixed head or chin rest. The device adjusts to ambient light levels using light sensors. The outcome measure was the binocular VF mean deviation (Bin-VF-MD).

All visual functions were performed using BCVA except for Bin-VF where near refractive correction is not mandatory as long as the VA < 0.1 logMAR.

### Statistical Analysis

After testing for normal distribution (Shapiro-Wilk test) and for assumptions for repeated measures analysis of variance (RM-ANOVA), data were analyzed and presented accordingly. The analyses of standard clinical test outcomes were performed employing a 3-way RM-ANOVA with the factors TIME (pre and post), INTERVENTION type (MMI and UMI), and GROUP (GLA AND HC). In addition, data of the specific visual tests performed during the different TM walking conditions were analyzed using a 4-way RM-ANOVA with the factors: TIME, INTERVENTION, GROUP, as well as SPEED (S_0_], S_3.5_ and S_Self_) as detailed in Table 4. If a violation of sphericity was detected, the Greenhouse-Geisser correction was applied. In case of significant interactions or main effects, post-hoc tests with Sidak-correction were performed. The effect size partial eta-squared 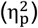 was calculated and interpreted as small (0.01-0.05), medium (0.06-0.13), and large effect (≥ 0.14) according to Cohen (1988). Effects were classified as significant if p ≤ 0.05 and relevant, if at least a medium effect size was observed. Data analysis was conducted in R (R Core Team, 2023) and SPSS (Statistical Package for the Social Sciences; IBM, Armonk, NY, USA).

## Results

Participants (18 GLA and 30 HC) were randomly assigned to either a MMI (GLA: n = 8, HC: n =12) or UMI group (GLA: n = 7; HC: n = 12). There was no age difference between groups (p = 0.276, Table 3). Three GLA and 6 HC had dropped out during the intervention period due to personal reasons. Baseline and demographic characteristics of the included participants are given in Table 3.

**Table. 3.**
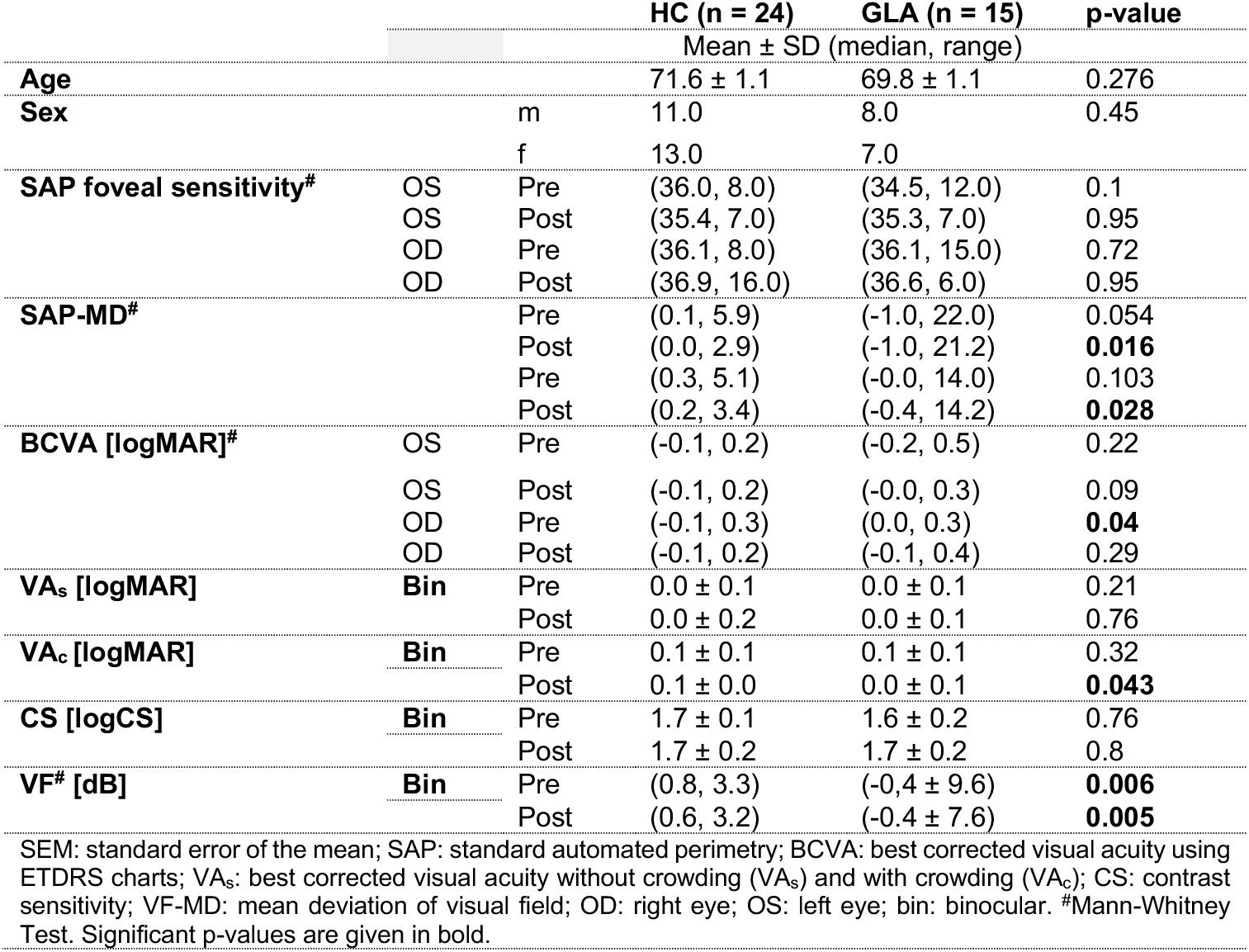
Patients demographic and clinical results before and after the 12-week interventions.

We next detailed the effects of the 12-week intervention on A) standard clinical tests and B) the TM based visual functional tests.

### A) Effect of the interventions on standard clinical tests

Main effects: We observed a significant TIME effect with regard to ETDRS BCVA 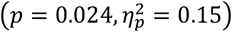, i.e., an improvement by 0.03 logMAR after the 12-week interventions. Interaction effects: There was a significant TIME x INTERVENTION interaction effect for the VF parameter ‘foveal sensitivity’ 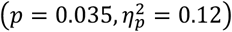. Post-hoc tests indicated an improvement of foveal mean sensitivities only for UMI, by 1.07 dB compared to the pre-intervention foveal sensitivities (35.2 dB), *p* = 0.024, (Table 4A).

**Table 4.** Overview of statistical outcomes of the RM-ANOVAs including all significant effects of SPEED, TIME, and INTERVENTION type on visual function parameters for the study GROUPs.

| Effect | F | p-value | partial eta-squared |
| --- | --- | --- | --- |
| **(A) Clinical parameters** |  |  |  |
| **ETDRS BCVA** |  |  |  |
| TIME | 1, 33 = 5.6 | 0.024 | 0.15 |
| **Foveal sensitivity [dB]** |  |  |  |
| TIME X INTERVENTION type | 1, 34 = 4.6 | 0.039 | 0.12 |
| **(B) Dynamic visual function** |  |  |  |
| **VAs [logMAR]** |  |  |  |
| SPEED | 2, 70 = 19.0 | < 0.001 | 0.35 |
| TIME x SPEED | 2, 70 = 3.1 | 0.052 | 0.08 |
| **VAc [logMAR]** |  |  |  |
| SPEED | 2, 70 = 9.1 | < 0.001 | 0.21 |
| **CS [logCS]** |  |  |  |
| TIME | 1, 35 = 3.8 | 0.059 | 0.10 |
| **Bin-VF mean deviations [dB]** |  |  |  |
| SPEED | 2, 66 = 124.9 | < 0.001 | 0.79 |
| GROUP | 1, 33 = 6.49 | 0.016 | 0.16 |
| TIME x GROUP | 1, 33 = 3.6 | 0.066 | 0.10 |
| INTERVENTION type x TIME x SPEED | 2, 66 = 2.34 | 0.105 | 0.07 |
| F: F-value of repeated measures ANOVA. For abbreviations, see Table 3. As shown in methods we used either $p < 0.05$ or effect size of 0.06 as a cutoff for any significant. For further details, refer to results. | | | |

### B) Effects of the interventions on TM walking visual function

#### (1) TM walking SPEED effects (S_0_ vs S_3.5_ and S_Self_)

Some of the results of the pre-intervention data-set have previously been reported by Beyer et al. (2024). Of note, the present study analyzed data of a smaller sample due to dropouts during the intervention period (GLA: 3, HC: 6). Main effects (significant effects detailed in Table 4B): For VA_s_, the mean BCVA static (mean ± SD: -0.015 ± 0.11 logMAR) was better than dynamic readings (mean ΔS_0_-S_3.5_ [ΔS_0_-S_Self_] ± SD: −0.03 ± 0.04, *p* < 0.001 [−.0.05 ± 0.04, *p* = 0.001]), see Figure 2A. Participants also had better VA_C_ at S_0_ (0.031 ± 0.09 logMAR) than S_3.5_ [S_Self_] (mean S_0_-S_3.5_ [S_0_-S_3.5_] difference ± SD: −0.02 ± 0.05, *p* = 0.10 [−0.04 ± 0.06, *p* < 0.001)]). Bin-VF were reduced during dynamic compared to static viewing (S_0_: 0.12 ± 01.83 dB) by a mean deviation loss of − 0.9 ± 0.44 dB during both S_3.5_ and S_Self_, *p* < 0.001, see Figure 2C. For CS there was no effect of speed. (comparable CS across different speeds with an average of 1.65 ± 0.16 logCS), see Figure 2B.

**Figure 2.**
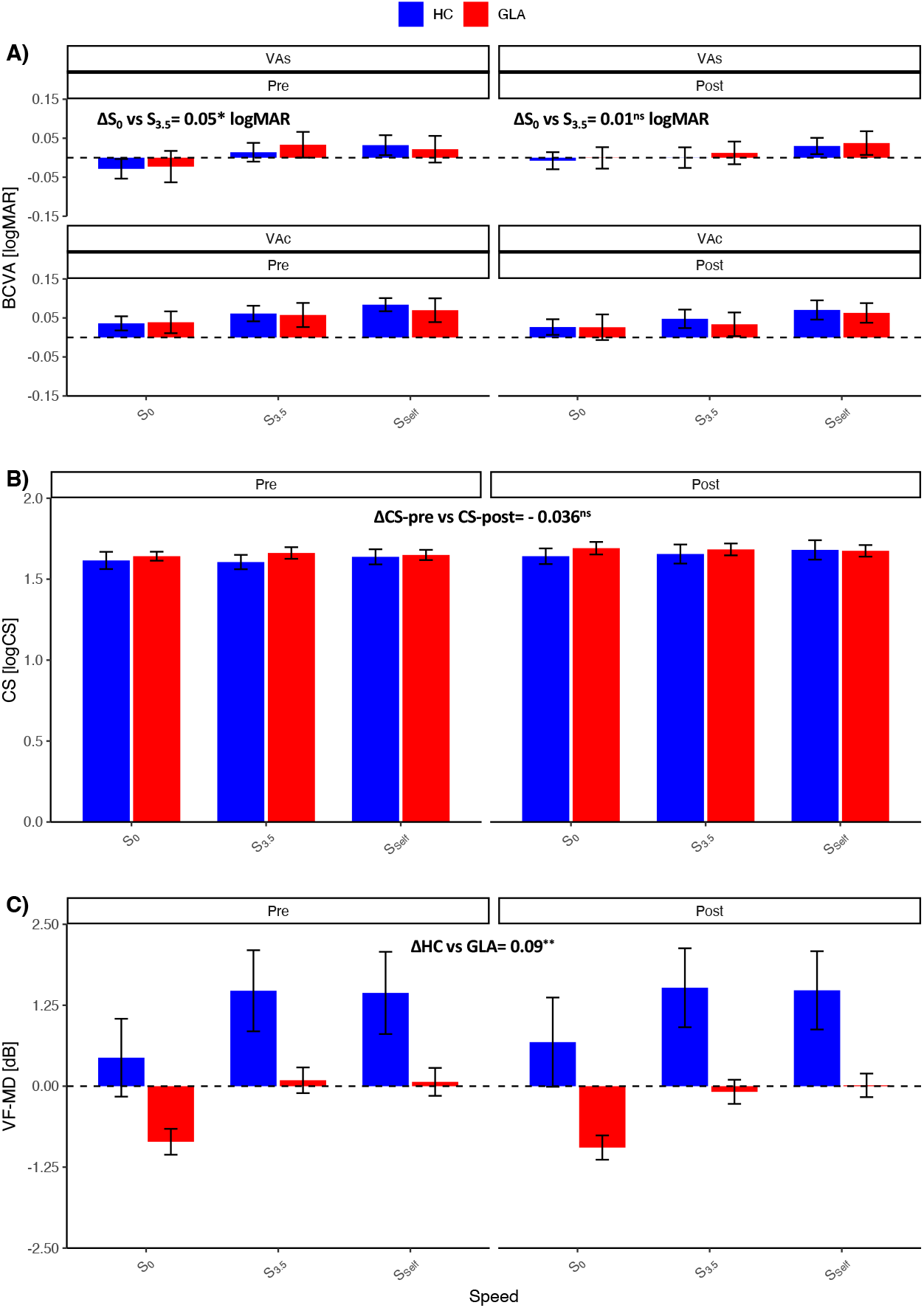
Visual performance before (pre) and after (post) the 12-week interventions (MMI & UMI) for 3 TM walking speeds. A) TIME (pre- and post-intervention) effects across different TM walking speeds for best corrected visual acuity with (VA_c_, upper panel) and without (VA_s_, lower panel) crowding. B) TIME (pre- and post-intervention) effects on contrast sensitivity (CS). C) TIME (pre- and post-intervention) effects on binocular visual field mean deviations (VF-MD) in decibels (dB). INTERVENTION types are combined for these effects. For statistics, see Table 4. Dotted line indicates zero-read-out. Intervention types’ effects are pooled due to lack of significant differences.

#### (2) TIME effects

Overall, visual function did not differ significantly after the 12-week intervention. Main effects: We observed a non-significant small trend of CS improvement after intervention (by 0.04 ± 0.1 logCS, p = 0.059, 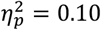). Interaction effects: For VAs, there was a non-significant trend of TIME x SPEED interaction effect, p = 0.052, η^2^ = 0.08;

#### (3) INTERVENTION effects (MMI vs UMI)

The intervention effects were not differentially affected by the two intervention types (MMI or UMI). Main and interaction effects: There were no significant effects.

#### (4) GROUP effects (GLA vs HC)

GROUP main effect: Only for Bin-VF, VF mean deviations were reduced for GLA, as expected from the pathology, −1.20 ± 2.4 dB than HC, 0.25 ± 0.91 dB, p = 0.016. Interaction effects: None detected.

## Discussion

The present study investigated the effect of MMI and UMI on visual function (i) tested with clinical parameters and (ii) during TM walking at different speeds. While trends were evident, the effect of the 12-week interventions on visual function were small and largely independent of group and intervention type.

### Intervention effects on visual performance

The observed intervention effects were marginal and reached (weak) significance only for the standard clinical measures of BCVA and foveal luminance sensitivity. This absence of strong effects is likely due to the early stage-nature of most of the included patients. Accordingly, Lee et al., investigating more advanced glaucoma cases, reported a 10% improvement of glaucomatous VF following 1-week of moderate-vigorous physical exercise (Lee et al., 2019). Even in healthy athletes, exercise, namely ocular-motor, has beneficial effects on dynamic visual acuity (Minoonejad et al., 2019). These positive effects are not confined to visual function, but also extended to cerebral health and cognition as shown for healthy elderly (Intzandt et al., 2021). In patients with mild cognitive impairment, aerobic and resistance raining has been shown to improve measures of brain function and structure (Huang et al., 2022). Thereby, the higher cognitive function seems independent of the intervention type (Intzandt et al., 2021) meaning that both physical and cognitive training have beneficial effects. In fact, these interventions are of high relevance to diseases, such as glaucoma and Alzheimer’s, upon validation in larger cohorts.

Several factors might explain the lack of relevant positive effects from our interventions on visual function readouts, including the low sample size and the early stage of disease progression in glaucoma patients as detailed below. In fact, the intervention effects in early glaucoma might be better captured by other outcomes related to visuo-cognitive-motor skills, including kinematic measures during physical locomotion, rather than basic visual function.

### Limitations and Outlook

When interpreting the results of the present pilot study, several limitations have to be considered. First, the sample size was small mainly due to the impact of the COVID-19 pandemic on the recruitment and regular study/intervention participation, as reflect in the “pilot study” nature of our report. Second, most glaucoma patients were in the early stages of glaucoma, which may have resulted in the small magnitude of the observed effects of the interventions as one might have expected in the later stages. This is confirmed by our previous findings (Beyer et al., 2024) that healthy elderly and early glaucoma participants have comparable visual function loss during TM walking. Hence, to realistically capture intervention impacts, in future studies more advanced glaucoma stages should be included where functional deficits are larger and might benefit from our training programs.

## Conclusion

In summary, while the intervention approach of the present study established a foundation for a novel movement-related intervention regime for glaucoma, the effect of the 12-week interventions specifically on visual function were small. Based on the data of the present study, future trials should recruit a larger sample size and participants with more severe glaucoma stages in order to identify the effects MMI und UMI on visual function to prevent glaucoma-induced loss of quality of life.

## Data Availability

All data produced in the present study are available upon reasonable request to the authors.

## Acknowledgements

We thank the study participants for their support of the study and gratefully acknowledge support by the German Research Foundation (DFG; Project: 423926179; HO-2002/20-1 & SCHE 1584/5-1). There was no rule of the funders in planning, conducting or reporting the current study. We acknowledge support by the Open Access Publication fund of the medical faculty of the Otto-von-Guericke-University Magdeburg.

## Notes

### Competing Interest Statement

The authors have declared no competing interest.

### Clinical Trial

The trial was registered at the German clinical trials register (DRKS00022519).

### Author Declarations

The experimental protocol was approved by the Ethics Committee of the Otto-von-Guericke University of Magdeburg in Germany (registration number: 32/18) and all procedures were in line with the Declaration of Helsinki on experiments on human beings

